# Negotiating the Social Self in Chronic Spinal Pain: A Qualitative Study

**DOI:** 10.64898/2026.11.06.26367446

**Authors:** Yvan Sonjon, Alice Charriau, Dihia Mouhoub, Jean-Baptiste Van der Henst, Eric Chabanat

## Abstract

**Purpose:** This study examined how people living with chronic spinal pain experience, interpret and navigate social representations of their condition and the interactions these generate across clinical, occupational, and personal settings.

**Materials and Methods:** Thirty semi-structured interviews were analysed using an approach informed by constructivist grounded theory, through initial, focused and theoretical coding.

**Results:** Five categories emerged, capturing how social representations and relational dynamics threatened the social self and prompted deliberate strategies for restoring agency. Theoretical coding identified a central process (maintaining and rebuilding the social self), describing how participants navigated these threats across social contexts. Notably, approximately half the participants reproduced, without revision, representations of chronic pain typical of the general population, directing judgements of simulation or inadequate effort at others with the condition. These strategies primarily aimed to restore agency and maintain a viable social self, rather than to reduce pain.

**Conclusions:** These findings describe one process through which social environments shape the experience of chronic spinal pain by influencing how the social self is continually negotiated. Recognising these processes may help rehabilitation professionals attend to the social dimensions of chronic pain.

**Implications for rehabilitation:**

- Rehabilitation professionals should recognise that people living with chronic spinal pain often face a substantial and largely invisible burden beyond pain management itself. Continually demonstrating that their pain is real and legitimate can place an additional cognitive and relational burden on patients. Allowing sufficient space in clinical encounters for patients to describe these experiences may help clinicians identify the social difficulties associated with chronic pain and how they affect daily life and participation.
- Shared experience of chronic pain should not be assumed to produce greater empathy toward others in similar situations. Approximately half the participants in this study continued to apply common social representations of chronic pain to other people with the condition. Therapeutic patient education and peer-based programmes should account for this possibility and actively address the representational frameworks participants bring to these settings.
- Rehabilitation assessment should include patients’ experiences of support, disbelief, paternalism, and changing expectations across healthcare, workplace, and personal settings, as these interactions may shape perceived credibility, control and self-efficacy.
- Clinical training in chronic pain rehabilitation should address both explicit delegitimation and more automatic forms of stereotyping, including how assumptions about what a person in pain should look like or do can influence clinical judgement and communication.

## Introduction

Chronic spinal pain, including non-specific low back and neck pain, presents a persistent paradox. For many people, the severity of the pain experience exceeds what biological and radiological investigations can explain (Brodal, 2017; Maher et al., 2017). Moreover, even the variables that do predict the condition’s evolution point less to symptom intensity than to psychosocial and cognitive factors (Mehling et al., 2015). Neuroimaging studies suggest that the transition to chronic pain involves a gradual shift in brain activity from nociceptive circuits towards regions involved in emotional processing (Hashmi et al., 2013), indicating that pain is not reducible to a passive response to sensory input. It is an experience shaped by the integration of sensory signals, cognitive and affective states, and the social and relational contexts in which it is interpreted and communicated (Brodal, 2017; Melzack, 2001). Sullivan et al. (2023) argue that the processes sustaining chronic pain are “*transdermal*”, having causes both inside and outside the body, and that psychological and social factors contribute to pain itself rather than merely modifying a biologically produced experience.

Social factors, including healthcare relationships, occupational conditions, and the representations that structure how pain is evaluated and communicated, are increasingly recognised as shaping the experience and course of chronic pain (Sullivan et al., 2023; Karran et al., 2020). However, how these influences are studied does not always capture the relationships and interactions through which they are experienced. In a previous scoping review, we identified 28 categories of social factors across six domains associated with the development of chronic low back pain (LBP) (Sonjon et al., 2025). Most of the included studies relied on quantitative designs in which social factors were operationalised through measurable individual characteristics or predefined categories, and were sometimes used as adjustment variables rather than examined as direct contributors to pain. These approaches provide limited access to the relationships and interactions through which social influences are experienced in everyday life. The literature thus tends to frame individuals through characteristics and categories rather than through the relationships and interactions in which their experience of chronic pain occurs. Recent critical work on the biopsychosocial model in LBP care has similarly shown that social dimensions are often only briefly addressed, with limited attention to interpersonal, cultural, and institutional power dynamics (Cormack et al., 2022; Mescouto et al., 2022). Our review highlighted an important gap, namely that patients’ experiences within healthcare settings, including the relational dynamics of clinical encounters, were underrepresented in the literature.

One relational process that remains less often documented concerns how people with chronic pain are evaluated by others, including the credibility granted to their account of pain, the extent to which they are suspected of exaggeration, and whether their suffering is regarded as worthy of recognition (De Ruddere et al., 2013; De Ruddere and Craig, 2016). Thus, within their social environment, people living with chronic spinal pain encounter widely shared, often reductive, representations of pain and of those who experience it. Chronic pain is largely invisible to others, and when its severity cannot be accounted for by demonstrable tissue damage, its legitimacy may be called into question (De Ruddere and Craig, 2016). Against this background, in clinical, occupational, and family settings, people may encounter suspicions of exaggeration, malingering, lack of will, or secondary gain (De Ruddere and Craig, 2016; Perugino et al., 2022; Zajacova et al., 2021). Such stereotypes about people with chronic pain shape how behaviours are interpreted and may influence the credibility afforded to a person’s account (García-Martínez et al., 2022). They are not necessarily expressed explicitly or mobilised deliberately, as evaluations may partly rely on automatic processes of social categorisation and implicit biases that can affect interactions and clinical decision-making (Amodio, 2014; Puddifoot, 2019). These representations may also be internalised by people living with chronic pain, shaping how they interpret their own situation (Perugino et al., 2022; Waugh et al., 2014).

These processes of social evaluation can be examined through the lens of social cognition, understood as the set of processes through which individuals perceive, interpret, and respond to social information in their interactions with others (Arioli et al., 2018; Lieberman, 2007). Social interactions also shape self-representations, including how individuals perceive their identity and social position in relation to others (Markus and Wurf, 1987). In chronic pain, Karos et al. (2018) propose that pain can threaten the social self by shifting control away from the person with pain, disrupting belonging through exclusion or invalidation, and giving rise to experiences of injustice. This last dimension may have consequences for pain itself. Among people with nonspecific chronic LBP, Penn et al. (2020) found that perceived injustice helped explain the association between chronic pain stigma and movement-evoked pain severity. Taken together, this work suggests that the social environment actively shapes pain experience through the interactions and relationships in which pain is lived. Attending to the accounts of people living with chronic spinal pain provides insight into how these processes are experienced and interpreted across interactions ranging from close relationships to healthcare systems and institutional settings.

The clinical relevance of psychosocial dimensions is well established, through their associations with quality of life in chronic LBP and the benefits of care that takes psychosocial obstacles to recovery into account (Agnus Tom et al., 2022; Hill et al., 2011). What remains less well documented is how these social situations and interactions are experienced by people living with chronic pain. Qualitative research has begun to document what matters to people within chronic pain care pathways (Franklin et al., 2016). Building on this work, the present study focuses on how representations of chronic pain, whether held by others or by people living with the condition themselves, are encountered in everyday relationships and institutional settings. The objective of this study was to explore, through participants’ own accounts, how people living with chronic spinal pain interpret the influence of these representations and interactions on their daily life, their pain experience and its evolution, and their healthcare pathway.

## Materials and methods

### Design

This study used an inductive approach and a qualitative design informed by the principles of constructivist grounded theory, as developed by Charmaz (2017). Within this paradigm, categories and theory are not considered to pre-exist the data. They are constructed through the interaction between researchers and participants, from situated knowledge embedded in a social environment. This epistemological position concerns the way in which the analysis constructs its categories, not the nature of pain or cognition. This approach was chosen because it makes it possible to explore, from the data themselves, how social representations and interactions shape the social experience of chronic pain, and to develop a theoretical model grounded in the experience of those concerned, one that can then be brought into dialogue with concepts and findings from the scientific literature. Data were collected through individual semi-structured interviews, using a guide that was stabilised after approximately the first ten interviews. Although early memos accompanied data collection, formal coding began once all interviews were complete. The study was therefore informed by constructivist grounded theory, rather than designed as a full application of it: its constructivist epistemology and analytic procedures, described in the Data analysis section, were retained, whereas the fully iterative and concurrent logic of theoretical sampling (i.e., directing data collection according to emerging theoretical categories) was not applied. Consistent with contemporary constructivist grounded theory guidance (Charmaz and Thornberg, 2021), the study retained an emphasis on reflexivity, transparency, and an open, non-deductive engagement with the existing literature.

### Researcher reflexivity

Individual interviews were conducted by the lead investigator, YS, and by AC, with YS conducting the majority of them. YS is a man, a physiotherapist and a doctoral researcher in cognitive neuroscience, with ten years of clinical experience in physiotherapy and prior academic experience in research on chronic pain and social cognition. During recruitment and at the beginning of each interview, he introduced himself to participants as a doctoral researcher in cognitive neuroscience, rather than in a clinical capacity, and explained the general topic of the study, which concerned representations and interactions around chronic pain. AC is a woman, a physiotherapist and a second-year Master’s student in rehabilitation research, with prior clinical experience in physiotherapy. During recruitment and at the beginning of each interview, she introduced herself to participants as a second-year Master’s student conducting research in the field of rehabilitation. No prior relationship existed between the investigators and the participants before the first contact made to arrange the interview. In line with Charmaz and Thornberg’s requirement for *“methodological self-consciousness”* (Charmaz and Thornberg, 2021), which involves making one’s assumptions explicit rather than setting them aside, the investigators’ clinical background as physiotherapists was considered both a resource and a potential source of bias. This background gave them familiarity with people living with chronic pain and with how chronic pain is typically framed in rehabilitation settings, which likely facilitated rapport during interviews and helped them follow the healthcare contexts participants described. It also risked steering interpretation toward clinically familiar categories, such as biomedical explanations of pain or standard descriptions of fear-avoidance, rather than the framings participants used themselves. YS’s prior academic work on chronic pain and social cognition added a further layer of vigilance, as this background may have sensitised the analysis to certain dimensions of the material, including representations, social judgments and interactions. Initial coding of a subsample of 12 participants was also conducted independently by DM, a woman, a general practitioner and a second-year Master’s student in disability and cognition, before being compared with the initial coding produced by YS on the same subsample. This comparison provided an external perspective on whether the emerging codes reflected participants’ accounts rather than the lead investigator’s prior clinical or theoretical assumptions. Theoretical coding leading to the identification of the central process was then developed independently by YS, JBVH and EC, before their respective analyses were compared and discussed in order to reach a shared central process. EC is a man, an associate professor and a psychologist. JBVH is a man, a CNRS research director in social cognition. The diversity of these clinical, methodological and theoretical backgrounds strengthened the grounding of the interpretation in participants’ accounts by bringing the lead investigator’s reading into discussion with independent perspectives.

### Study participants

Participants were recruited from several Pain Evaluation and Treatment Centres (Centres d’Évaluation et de Traitement de la Douleur, CETD) in the Auvergne-Rhône-Alpes region, France. The centres are not named in order to preserve the anonymity of the recruitment context. These centres correspond to a tertiary level of care for chronic pain, generally reached after a prolonged care pathway. This setting was chosen to recruit participants with substantial first-hand experience of living with chronic spinal pain and of navigating the healthcare system in relation to it.

Eligible participants were adults aged 18 years or older with a diagnosis of chronic spinal pain, either chronic non-specific LBP or chronic neck pain, persisting for more than three months despite treatment (Haute Autorité de Santé, 2019), with or without peripheral radiation. Patients were not included if their spinal pain was attributable to a tumoural or infectious cause, or to vertebral fracture or dislocation. They were also not included when unexplored or newly occurring warning signs suggested an underlying pathology requiring specific or urgent care, such as unexplained weight loss, a history of cancer associated with fever, thoracic pain, fever, or deterioration in general health status. These criteria were based on a subset of the red flags described by the Haute Autorité de Santé (HAS) for the assessment of common LBP (Haute Autorité de Santé, 2019), transposed to chronic neck pain. Radiating pain, neuropathic components, or known neurological symptoms were not exclusion criteria when they had already been investigated, monitored, or managed by the pain physician. In all cases, participation depended on prior medical screening performed by the pain physician managing the patient, on the basis of the medical diagnosis established during a scheduled consultation.

### Sampling

A purposive sampling strategy was used, combining an a priori target sample size with an internal diversification strategy intended to capture a diversity of experiences within a homogeneous population.

The target sample size of 30 participants was set before data collection began. Marshall et al. recommend that grounded theory studies generally include between 20 and 30 interviews, and report that, in their review, studies exceeding this range do not show evidence of greater scholarly impact (Marshall et al., 2013). This sample size was also considered consistent with the principle of *information power*, according to which sample adequacy depends on the aim of the study, the specificity of the participants, the use of existing theoretical frameworks, the quality of the dialogue, and the analytic strategy (Malterud et al., 2016). It also seemed ethically preferable not to continue recruitment beyond a number of participants likely to provide sufficient information in relation to the study aim. Because data collection and analysis were conducted sequentially (see *Design*), saturation was not assessed iteratively during recruitment. The sample size of 30 participants was therefore set in advance as a criterion of sample adequacy, rather than as the result of theoretical saturation assessed during the course of analysis.

The homogeneity criterion defining the study population was the presence of chronic spinal pain, including chronic non-specific LBP and chronic neck pain. Internal diversification aimed, as far as possible, to include a range of participants within this population, irrespective of their statistical frequency in the clinical population. The dimensions sought included pain location (low back or neck pain), pain duration, age, gender, socio-economic status, and area of residence. This strategy aimed to enrich the range of experiences collected and to support comparison of internal variations within the phenomenon under study, namely chronic spinal pain (Malterud et al., 2016). Chronic LBP and chronic neck pain were therefore treated as variations of the same phenomenon, examined through the same analytic framework, rather than as two populations requiring separate explanatory frameworks.

### Recruitment

Recruitment and inclusion of participants took place between March 2024 and February 2025. Potential participants were identified by physicians from the CETD during scheduled consultations, based on the medical diagnosis and the eligibility criteria described above. Eligible patients received oral information and a written information sheet from their physician. Patients who agreed to be contacted were then approached by the lead investigator (YS) or by the investigator (AC), by telephone or email, to arrange an interview. Recruitment was also supported by information posters displayed in the waiting rooms of the CETD. All patients who expressed interest in the study and were contacted by the investigators took part in the semi-structured interview planned as part of the study. Participants did not receive any compensation for their participation.

At the beginning of each interview, information about the study and the interview procedure was repeated to the participant. Participants were able to ask any questions about the study before the interview began. Before recording, they also signed a form authorising the recording and processing of their voice. Each participant was reminded that they could take a break during the interview, decline to answer a question, choose not to discuss a topic, stop the interview, or withdraw their consent to participate and to the use of their data at any time, including after the interview had ended. Participants could also request that specific passages from the interview be removed before data analysis.

### Data collection

Data were collected through individual semi-structured interviews conducted by YS and AC. Each participant took part in a single interview session, with an indicative duration of approximately one hour. Interviews took place either in a dedicated room at the CRNL (Lyon Neuroscience Research Centre, Bron) or on the premises of the participating CETD. A secure videoconferencing option (BigBlueButton) was available at the participant’s request and in accordance with lockdown requirements in cases of suspected or confirmed COVID-19 infection, but no interviews were ultimately conducted by videoconference. No person other than the participant and the investigator was present during the interviews, and no repeat interviews were conducted.

At the beginning of each interview, the investigator explained the general aim of the study, stated that there were no right or wrong answers, and reminded the participant that they were free not to answer a question or to return to it later. Before recording began, the participant’s agreement was confirmed and they were informed that the recording was starting. Interviews were recorded using a Philips VoiceTracer digital recorder. A pseudonymisation code, generated for each participant before the interview, was read aloud at the beginning of the audio recording.

The semi-structured interview guide was developed in collaboration with researchers in sociology and anthropology (see *Acknowledgements*) and is provided as supplementary material. In keeping with the inductive nature of the study, the initial version of the guide included a dedicated theme on the history and subjective experience of pain, and explored the participant’s healthcare pathway through two separate questions, one addressing diagnosis and consultations, the other addressing the experience of being accompanied by healthcare professionals and by personal or professional networks. Following a preliminary review of the material collected from approximately the first ten interviews, conducted independently of formal coding (see *Design*), the guide was revised. The theme addressing the history and subjective experience of pain was no longer retained as a dedicated theme, in order to refocus the interview on social representations and interactions around chronic pain rather than on the phenomenological experience of pain itself. The two questions on the healthcare pathway were merged into a single question. A new question exploring the participant’s sense of proximity or belonging with other people living with chronic pain was also added. The remaining domains of the interview guide were unchanged. This revised version was used for the majority of interviews. The final version of the interview guide is provided as supplementary material 1 on Zenodo (https://doi.org/10.5281/zenodo.23963293) (in French, the language in which interviews were conducted).

The guide began with questions about the participant’s biography and, for the majority of interviews, the place of pain in their daily life, in order to better understand their context and create a climate of trust before addressing the central topics of the study. Interviews then explored the participant’s representations of chronic pain and of people living with it, the representations they attributed to healthcare professionals and to their personal and professional networks, and the relational and bodily consequences of these representations for their daily life, their pain, and their healthcare pathway. The guide also addressed the participant’s relationship to other people living with chronic pain, including any sense of proximity, belonging, or mutual recognition. Demographic information, including age, gender, family situation, socio-economic status, education level, area of residence, mode of transport, lifestyle and leisure activities, frequency of healthcare consultations, pain duration, and current treatments, was collected at the end of the interview when it had not already been discussed. This choice aimed to avoid beginning the interview with a series of closed questions that could have elicited socially expected answers or conditioned the rest of the exchange.

Once the interview was completed, the participant was asked whether they wished to add anything they considered important for this research before the recording was stopped. After the recording had been stopped, time was set aside for debriefing with the participant. Interviews were audio-recorded and then transcribed verbatim for analysis. Transcripts were not returned to participants for review or correction. This choice was made for ethical reasons: most participants did not have access to a secure email account through which transcripts could have been transmitted safely, and several participants experienced mobility difficulties related to their chronic pain, with some relying on medically equipped transport services that could not be arranged for an additional visit to the research sites. Requiring participants to travel again for this purpose was considered to impose an unjustified burden.

### Data analysis

Interview transcription was carried out by YS, AC and DM. Once transcription was completed, the audio recordings were destroyed, in accordance with what had been agreed with the Data Protection Officer (DPO) during the ethical assessment of the study, in order to protect participants’ pseudonymity as far as possible and to minimise the existence of identifiable personal data. The transcribed verbatim accounts were managed and coded using NVivo software (QSR International), release 1.7.2 (1560). The final codebook, including category definitions, hierarchical structure, and the number of coded interviews and segments per code, is provided as supplementary material 2 on Zenodo (https://doi.org/10.5281/zenodo.23963293).

The analysis followed the principles of constructivist grounded theory as described by Charmaz (2017). Data were analysed through initial coding, begun independently by DM and YS on approximately twelve interviews, then continued by YS alone after comparison of the emerging codes. During this phase, segments of the transcripts were named using active, process-oriented language. Several phases of revision took place after the analysis of 3, 7, 18, 25 and then 30 interviews. Codes were merged when they catalogued similar phenomena, deleted and redistributed to other codes when they were not significant in the corpus, or refined in their wording and definition.

Focused coding began halfway through the initial coding of the interviews and was conducted by YS alone. During this stage, the most significant codes were systematically compared, refined and developed into analytic categories. This step made it possible to structure a hierarchy between categories, codes and subcodes. Memo-writing accompanied the whole process of initial and focused coding.

Once initial and focused coding of the 30 interviews had been completed, theoretical coding began, with the aim of identifying a central process accounting for the relationships between categories. This stage was conducted independently by YS, JBVH and EC, each developing their own central process. The three analyses were then compared during joint meetings, in order to discuss their similarities and differences, until a single central process was agreed by the three authors and presented in *Results*.

Although sensitising concepts from the literature on social cognition and chronic pain informed the interpretive lens brought to the data, the categories were developed from participants’ accounts and remained grounded in them. In line with Charmaz and Thornberg’s recommendation for an open and non-deductive engagement with the existing literature (Charmaz and Thornberg, 2021), this literature was used as a sensitising resource rather than as a predetermined framework to be confirmed.

### Rigour and trustworthiness

Several strategies were used to support the trustworthiness of the analysis, understood through the criteria of credibility, dependability, confirmability and transferability discussed by Charmaz and Thornberg (2021) for grounded theory research. Although coding was not formally duplicated across the full corpus, initial coding was independently compared between DM and YS on a subsample of interviews. The central process emerging from theoretical coding was also developed independently by YS, JBVH and EC, before being reconciled through joint discussion. Transferability was supported by the internal diversification sampling strategy described in *Sampling*. Methodological self-consciousness regarding the lead investigator’s clinical background, maintained throughout the reflexive practices described above (see *Researcher reflexivity*), contributed to the confirmability of the analysis. Transcripts were not returned to participants for member-checking, for the ethical reasons described in *Data collection*; this is acknowledged as a limitation to the credibility of the findings. Reporting of this study was guided by the Standards for Reporting Qualitative Research (SRQR) (O’Brien et al., 2014) and the Consolidated Criteria for Reporting Qualitative Research (COREQ) (Tong et al., 2007).

### Ethical considerations

This study was conducted in full compliance with the principles of the Declaration of Helsinki. Ethical approval was obtained from the Research Ethics Committee of Lyon University (approval number: CER-UdL 2023-01-12-001). All participants provided written informed consent to participate in the study, for audio recording of the interviews until transcription, and for the publication of anonymized excerpts from their interviews. Data processing complied with the General Data Protection Regulation (EU Regulation 2016/679), in consultation with the institution’s Data Protection Officer.

## Results

### Sample characteristics

Thirty participants were included in the study. The mean age was 49.9 years (SD = 18.0, range 18-77), with 16 women (53.3%), 10 men (33.3%), and 4 non-binary individuals (13.3%). Pain was located in the lumbar region for 16 participants (53.3%), in the cervical region for 10 participants (33.3%), and in both regions for 4 participants (13.3%). The median pain duration was 8 years (range 0.5 to *≥*20). For nine participants (30.0%), pain duration exceeded 20 years but could not be determined more precisely; for descriptive purposes, these cases were recorded as *≥*20 years. Full sociodemographic and clinical characteristics are reported in table 1.

**Table 1.**
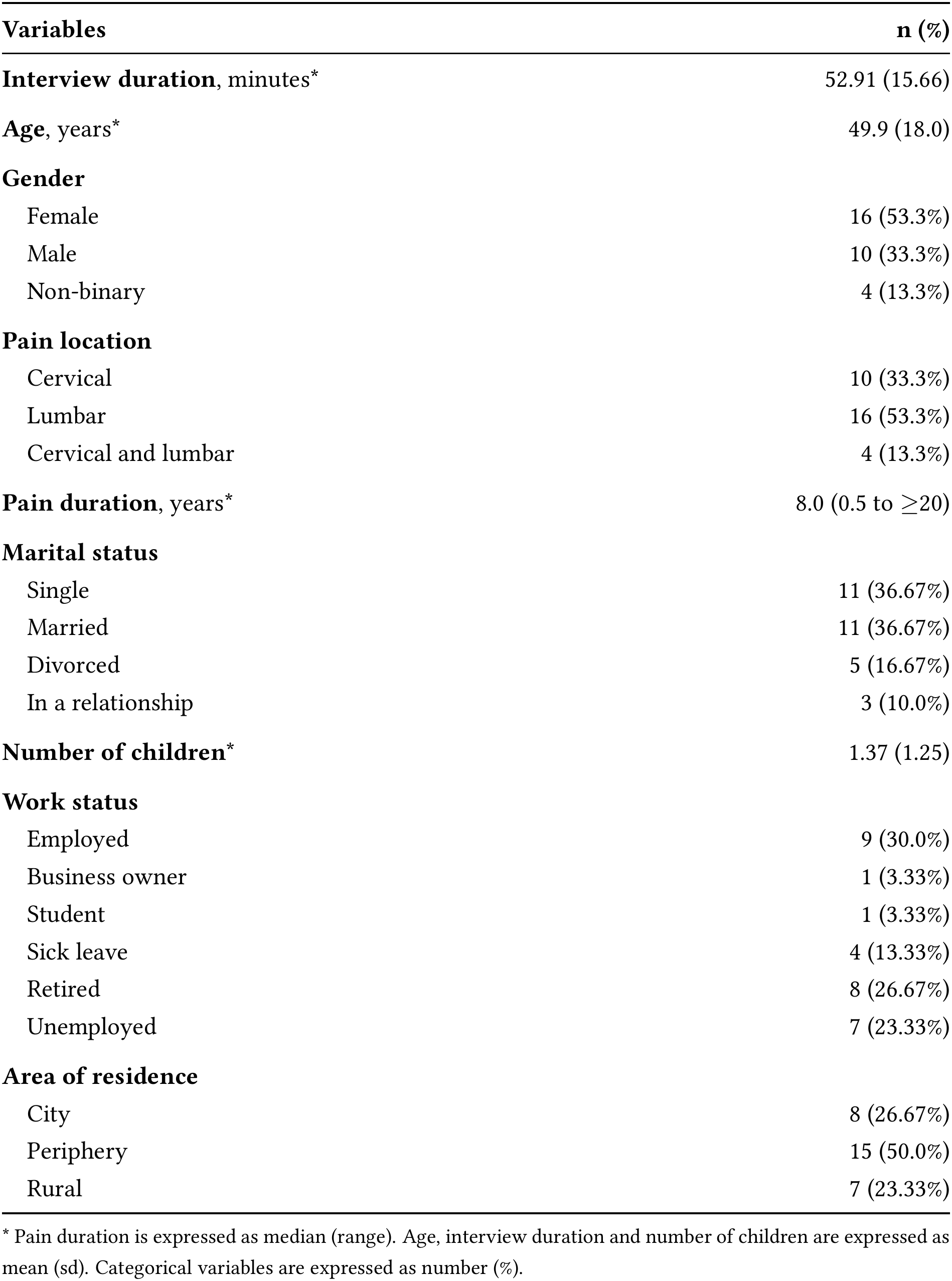
Demographic characteristics of the participants (*n* = 30)

### Categories from initial and focused coding

Initial and focused coding of the 30 interviews resulted in five categories. Each is presented below with its definition, internal structure, and illustrative quotations. The hierarchical organisation of the categories, codes, and sub-codes is shown in figure 1, and the complete codebook is provided as supplementary material 2 on Zenodo (https://doi.org/10.5281/zenodo.23963293). This structure reflects the organisation of codes within each category and does not imply any ordering or precedence among the five categories themselves. Participant quotations are identified by gender, age group, pain location, and pain duration to provide contextual information while preserving anonymity.

**Figure 1.**
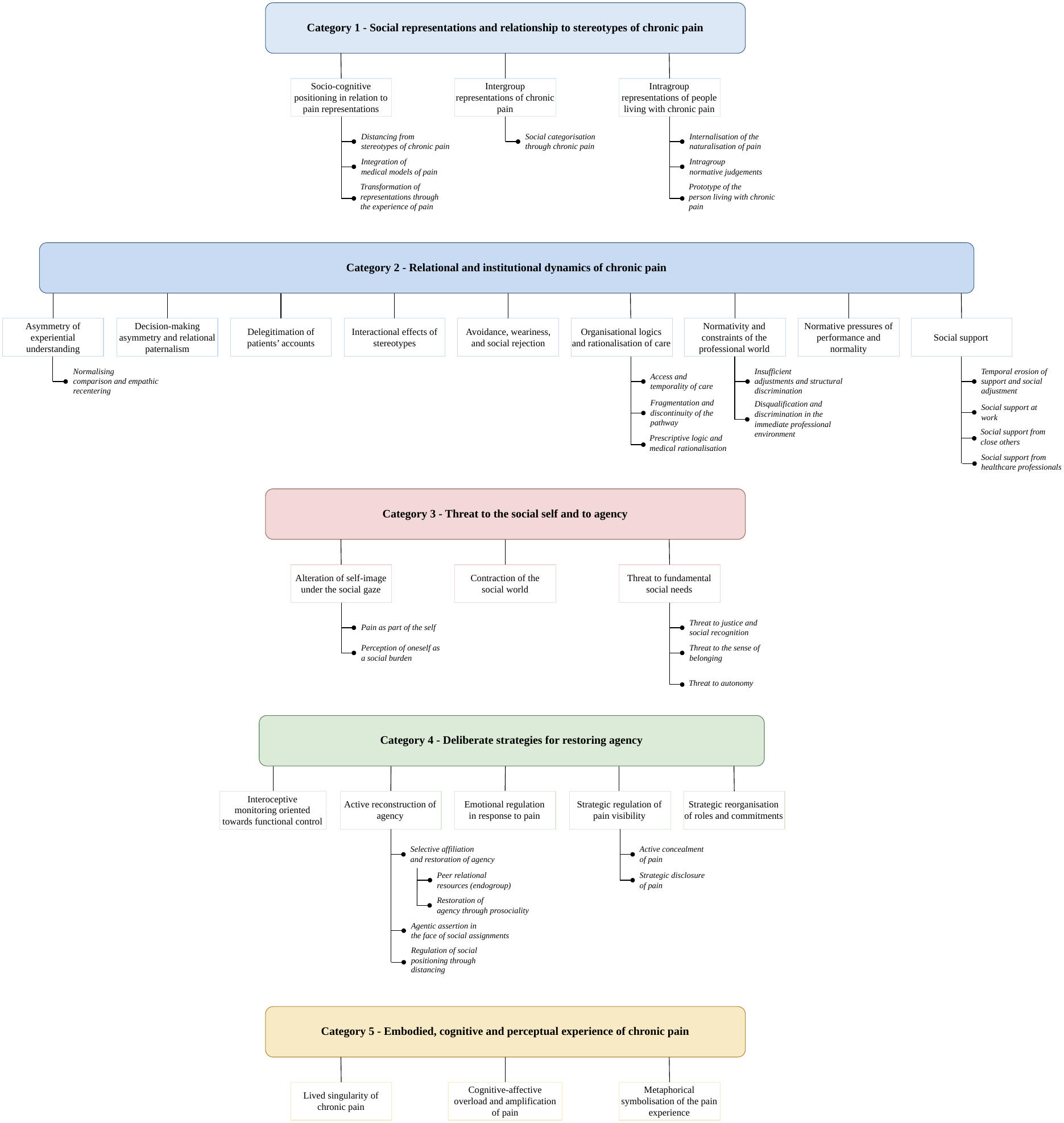
Hierarchical organisation of the emergent categories, codes and sub-codes. The figure illustrates the hierarchical organisation of the five emergent categories, their associated codes, and sub-codes identified through the qualitative analysis. Coloured boxes represent emergent categories, white boxes represent codes, and the indented text represents associated sub-codes. This figure presents the analytical structure of the findings, whereas the theoretical relationships between categories and the central process are illustrated in Figure 2.

#### Category 1: Social representations and relationship to stereotypes of chronic pain

This category brings together the ways participants described chronic pain and people living with it. Their accounts included bodily, behavioural and moral stereotypes, but also judgments about themselves, about other people with chronic pain, and about people without chronic pain. These representations mattered because they shaped how pain was evaluated in social situations and how interactions unfolded. Participants did not simply describe these representations; they also positioned themselves in relation to them, by contesting, transforming or internalising them in light of their own experience. One prominent pattern concerned how participants’ own experience of chronic pain related to the representations they held of other people living with the condition. The corpus revealed a clear divergence rather than a single pattern. For approximately half of the participants, lived experience left prior representations of other pain sufferers unchanged, leading them to validate stereotypes shared with the general population:

> “There are people who stop paying attention to their appearance, who, because of the pain, only wear what they call comfortable clothes, even though you can still look presentable, even quite stylish, and be comfortable at the same time. And then there is a kind of letting go, sometimes blaming the physiotherapist, saying, ‘He’s not capable because he can’t relieve my pain,’ or the doctor, ‘That one is useless.’ […] I think that sometimes it is easy to blame others, saying they are not capable of relieving me, whereas the pain is not our fault. You cannot say that, but it is still ours.” *Woman, 70–79yrs, Neck pain, ≥* 20*yrs*

For a comparable number of participants, the same lived experience prompted an explicit revision of previously held representations, leading them to invalidate stereotypes they had once shared:

> “Sometimes you have these stereotypes and think, you’re overreacting a bit, but when you find yourself in the same situation, you realise things differently. Looking back, I understood that these people, especially my mother, were actually in distress. And now, when things are difficult for her, I’m the first to support her, the first to say, I’m here. What do you need?” *Man, 30–39yrs, Low back pain, <* 2*yrs*

This shift could also take a more general form, as when one participant described being selfcentred and constantly attentive to his own body as “*something I would never have imagined*” (*Man, 60-69yrs, Low back pain, ≥* 20*yrs*). Participants also described a typical image of the person living with chronic pain, combining bodily signs, observable behaviours and psychological dispositions. This prototype could draw either on participants’ own experience as members of this group, or on representations they felt were projected onto them by people without chronic pain. In many accounts, these two sources were difficult to disentangle. One participant captured this composite figure:

> “Well, I think she’s an organised person. Very organised. She knows straight away what she can do and what she can’t do. She’s not someone who complains much, but she will say when it’s really too much, when she can’t go on. She’s someone who suffers. A lot. And it’s not only the physical pain. There’s the mental pain as well, and everything that comes with it.” *Man, 20–29yrs, Low back pain, 5–10yrs*

The image combined competence and restraint with a strong underlying sense of suffering. It therefore differed from other parts of the corpus, where the prototype was more often associated with resignation or irritability. These representations shaped what participants expected from social encounters before those encounters even took place.

#### Category 2: Relational and institutional dynamics of chronic pain

The social environment, from immediate interactions to organisational structures, shaped not only the recognition and legitimacy of chronic pain in social situations, but also participants’ understanding of their own experience and of their place within these interactions. These mechanisms took several forms, including interactional stereotypes, relational asymmetries, normative or institutional constraints, and supportive resources. These dynamics mattered because they affected whether participants felt believed, socially included, and able to make decisions about their own lives. One of the most consistently documented mechanisms was the delegitimation of participants’ accounts of their own pain, through psychologisation, suspicion of exaggeration, or explicit doubt. This differed from the more automatic effects of stereotyping, in which assumptions about what a person in pain should look like could shape an interaction without either party’s awareness. Delegitimation involved a more explicit questioning of what participants described, including whether their pain was real, how severe it was, or how it should be understood. For some participants, this had consequences that extended over years:

> “…I’ve been suffering with my back for several years. [Crying] I wasn’t taken seriously from the beginning, so I lost more than ten years. It’ll pass…” *Woman, 60–69yrs, Low back pain, ≥* 20*yrs, crying*
>
> “You have to get over the pain. […] It’s mental.’ It’s in your brain.’ It’s not physical pain, it’s mental pain.’ I heard that for a long time, from healthcare professionals, doctors, and family. […] When they can’t find anything physical, then it’s the brain.’ It’s a way out because they don’t know. An excuse. So yes, ‘it’s all in your head.’ […] And now we’ve had proof that it wasn’t just in my head. Maybe some of it was, but ninety-five percent of it was mechanical, physical.” *Man, 20–29yrs, Low back pain, 5–10yrs*

Against these threats to recognition, social support could act as a counterweight, but only when it preserved rather than substituted for the person’s autonomy:

> “My colleagues were always very considerate with me. Some of them knew I was unwell, and they would try to help, for example when we had to move boxes: No, don’t carry those, I’ll do it for you. Sometimes I still did it myself, but they were always considerate without going as far as saying, You’re disabled, you can’t do this or that.” *Woman, 60–69yrs, Low back pain, ≥* 20*yrs*

When support instead anticipated needs or imposed restrictions without being asked, it could reproduce, in a more benevolent register, the same threat to autonomy that delegitimation posed more overtly. Support was also not always stable over time; several participants described how the attention of those around them diminished gradually, even as their condition remained unchanged:

> “People around you understand. And sometimes they don’t. At first, they do understand, and then over time you get the impression that they forget. But my situation hasn’t changed. Over the past year and a half, I’ve maybe seen a few improvements, but they’re very small. The children found it difficult too, because they didn’t understand. […] It was so disabling.” *Woman, 50–59yrs, Neck and low back pain, <* 2*yrs*

Social support was therefore less a stable resource than something that required ongoing calibration, and could fail in two directions: by receding over time, or by crossing into decisions that were not its place to make. Beyond interpersonal relationships, participants also described how broader institutional environments, where social constraints became increasingly structural, could threaten their standing at work through norms of performance and productivity embedded in professional settings:

> “I feel like people are a bit like, You have back pain, you shouldn’t be doing this job. And when I notice little things related to that, I do feel a kind of threat: how long are they going to trust me? I feel like I don’t have the right to make a mistake, or to have a moment of pain, because otherwise I’ll be seen as a person in pain, someone who can’t swing a sledgehammer to put up the marquee, who can’t carry things. But actually, I really love carrying things. I love the physical side of my job.” *Woman, 20–29yrs, Low back pain, 2–5yrs*

The remaining dimensions of this category, including access to care, fragmentation of the care pathway, and discrimination in professional settings, are detailed in the codebook in supplementary materials.

#### Category 3: Threat to the social self and to agency

Where the social environment shaped the recognition and legitimacy of chronic pain from the outside, its consequences unfolded within the person. Chronic pain progressively weakened the social self through the internalisation of others’ real or anticipated gaze, the incorporation of pain into personal identity, the perception of oneself as a social burden, and the contraction of the relational world. These processes were endured rather than chosen, cumulatively altering participants’ perceived place, legitimacy, and sense of control over their own trajectory. Threats to three fundamental social needs, namely autonomy, belonging, and recognition, were documented across almost all thirty interviews. The most consistent finding was an alteration of self-image experienced as a break with a prior self. For several participants, chronic pain had not simply added a constraint to an otherwise continuous identity; it had produced a different person.

> “I feel like I’m not the same person anymore. There’s the sadness, of course, because I tell myself, Well, I’m going to be in pain for the rest of my life. That’s hard. But I also feel a little less… a little less joyful. I feel like it has changed my character, even though I’m someone who laughs all the time. Well, I try to. And I feel like I’ve become more withdrawn, more closed off, including with other people.” *Woman, 50–59yrs, Low back pain, 5–10yrs*

For many participants, this biographical rupture was accompanied by a parallel threat to autonomy, extending beyond functional limitation to affect the person’s sense of ownership over their own inner experience. One participant described how his colleagues’ repeated warnings about his pain began to shape his attention to his own bodily sensations:

> “So I pay more attention to it [the perception of pain] afterwards. Maybe they’re right, in the sense that I should be more aware of it in everyday life to avoid situations that cause pain. But because, for now, it doesn’t come from me, I experience it as a kind of loss of ownership over my own subjectivity.” *Non binary person, 20–29yrs, Low back pain, 2–5yrs*

This account illustrates a threat to autonomy that extended beyond the capacity to act, affecting the participant’s sense of authority over their own experience. The remaining dimensions of this category, including perception of oneself as a burden, contraction of the relational world, and threats to belonging and recognition, are detailed in the codebook in supplementary materials.

#### Category 4: Deliberate strategies for restoring agency

Where the preceding categories documented processes that were endured rather than directed, this category captured a qualitatively different register: the deliberate behavioural responses through which participants worked to restore their sense of agency and reconstitute a viable self. These responses operated across several domains. They ranged from sustained interoceptive monitoring and the careful calibration of physical effort to the strategic management of pain’s social presentation through decisions about when, with whom, and in what terms to disclose or conceal its existence. Participants also described asserting their own perspective against social and medical assignations, selectively affiliating with peers as sources of practical learning and recognition, and reorganising roles and commitments to preserve participation despite constraints. What distinguished these processes from coping in the conventional sense was not their variety but their orientation: they targeted the restoration of perceived control over one’s own experience and one’s place in the social world, not the reduction of pain itself. A recurrent strategy involved an active monitoring of bodily states, oriented towards functional control rather than pain relief. Participants described building, over time, a working expertise in reading their own signals:

> “You have to sort through the information that comes in, because when you live with chronic pain, you pay a lot of attention to your body. When I walk, am I going to be in pain? It’s like a sword of Damocles hanging over your head. The kind of pain I have gives no warning. It can start at any moment, in any situation. So you’re always on alert, knowing that it could start.” *Man, 60-69yrs, Low back pain, ≥* 20*yrs*

For several participants, monitoring increasingly involved anticipation. One participant illustrated this shift by describing a new pattern in her own thinking: before any physical movement, she would first ask herself whether she could do it, something she had not done before the onset of chronic pain. Participants therefore appeared to evaluate their bodily capacities before acting, based on what they had learned from previous experiences of pain. Another dimension concerned the deliberate management of pain’s social visibility. Participants made context-dependent decisions about what to reveal and what to conceal, to whom, and when. As one participant explained:

> “In general, I think people with chronic pain, you can’t see it on our faces. Even if sometimes I have to make it more obvious for certain doctors, it isn’t written on my face that I’m in pain everywhere. I’m not going to say every few seconds, Ah, I’m in pain, just to remind people. At some point, you learn to mask. People with pain mask, just like other people do. We try to be perceived well because we want to fit in. We don’t want to be isolated. You’re asking me about it now, so I’m telling you. But I’m not going to explain to my baker that I’m in pain everywhere.” *Man, 30-39yrs, Neck pain, 10-15yrs*

Concealment was only one aspect of this process. When disclosure was judged strategically useful, to negotiate workplace adjustments, to manage others’ expectations, or to establish the terms of a relationship, participants actively chose to make their pain visible on their own terms. Rather than representing opposite behaviours, concealment and disclosure formed complementary strategies through which participants sought to maintain control over their social participation. Against such social assignations, several participants described active selfassertion that went beyond information management. One participant described this effort:

> “I feel like the role I play in how people see this is to show that it happens, that it is possible to be young, active, athletic, to do physical work, and still have back pain. I don’t make it my everyday activism, but I do try to express that a lot. […] I think people often associate someone in pain with someone vulnerable, or someone in a position of weakness.” *Non binary person, 20–29yrs, Low back pain, 2–5yrs*

In doing so, participants challenged prevailing stereotypes while reclaiming a social identity threatened by chronic pain. Role reorganisation emerged as another important regulatory process. Participants described making deliberate choices about which activities, relationships and engagements to prioritise and which to reduce or relinquish, not as passive withdrawal but as a means of preserving a coherent sense of participation. For some, this reorganisation extended to a deeper revision of values and priorities, reshaping what it meant to occupy one’s roles as a worker, a parent, or a partner in ways that remained liveable. Finally, deliberate affiliation with peers who shared similar experiences differed from more general forms of social support. Participants described feeling understood without having to justify their pain, and sometimes used others’ experiences as a source of vicarious learning to adjust how they managed their own situation:

> “There was one person in particular who also had a lot of pain and who told me, ‘I’ve learned to manage it.’ I tried to draw inspiration from her. It brought me relief, first of all emotionally. I’m not saying that my chronic pain decreased, but it became more bearable.” *Woman, 70–79yrs, Neck and Low back pain, <* 2*yrs*

These interactions also reshaped participants’ understanding of chronic pain and those living with it, sometimes challenging previously held stereotypes and prototypes. Emotional regulation cut across each of these processes and is detailed in the codebook in supplementary materials.

#### Category 5: Embodied, cognitive and perceptual experience of chronic pain

This category specifies how the social processes described above also shaped the embodied experience of pain itself. It documents how participants attended to bodily signals, made sense of fluctuations, and tried to make an invisible experience communicable to others. Across participants’ accounts, chronic pain emerged as a deeply personal perceptual experience that resisted external quantification, saturated cognitive resources, and required considerable interpretative work to become communicable to others. Unlike the preceding categories, which traced how social interactions shaped participants’ sense of self from the outside, this category documented what that shaping produced within participants’ own perceptual and cognitive experience, and how that experience in turn altered their engagement with the social world. A recurrent finding concerned the fundamental incommensurability of chronic pain as a subjective experience. Participants consistently described a gap between externally observable markers and the intensity of what they experienced, making pain difficult to compare across people and difficult for others to fully grasp. One participant put this directly:

> “Pain can be on a different scale for each person, depending on what they feel. Sometimes that isn’t taken into account enough. You can say, for example, ‘Today my pain is seven out of ten,’ but my seven might be like twenty-eight for someone else. That’s an exaggerated example, but I think this is something people don’t always understand, including healthcare professionals, family, and people in everyday life.” *Man, 20–29yrs, Low back pain, 5–10yrs*

A second finding concerned the cognitive and affective weight of chronic pain. Participants described pain as persistently demanding attention and leaving less room for work, relationships and everyday activities. They also described a reciprocal relationship between pain and emotional states. Negative social interactions, including those described in Category 2, could increase stress and anxiety, which participants associated with greater pain. One participant described this directly:

> “Emotions really matter when it comes to managing pain. And when you keep taking hits all the time, of course it makes things worse. It increases stress, then anxiety, and that creates really unpleasant cycles where, in the end, it hurts even more.” *Woman, 20-29yrs, Neck pain, 5-10yrs*

Several participants also reported the converse: sustained engagement in an absorbing activity could temporarily attenuate perceived pain, only for it to return once the engagement ended. Such bidirectional dynamics between attentional allocation and pain modulation appeared across multiple accounts, pointing to a coupling between cognitive and perceptual processes that the social environment could activate in either direction. Present in a smaller part of the corpus, a further aspect concerned the cognitive work participants undertook to make chronic pain communicable. When direct description fell short, participants constructed metaphors to capture aspects of their experience that were otherwise difficult to put into words. One participant described the intensity of his pain on waking by telling his doctors that, every morning, he *“wakes up and looks for the debris of the house that fell on him during the night, as if someone had picked him up and thrown him to the ground”*. Another described the intimacy of the experience: pain, he said, *“is like a toothbrush, you don’t lend it to anyone. It’s something very intimate”*. These metaphors helped participants convey private perceptual experiences in terms that others could partially grasp. The limits of this communication connect with the asymmetries of understanding and delegitimation described in Category 2. The remaining dimensions of this category, including the progressive alteration of physical and cognitive capacity, the interplay between pain and sleep disturbance, and the existential weight of pain without prospect of resolution, are detailed in the codebook in supplementary materials.

Taken together, these categories describe more than a set of social experiences surrounding chronic pain. They show how chronic pain becomes socially interpreted, interactionally tested, embodied in perceptual and cognitive experience, and actively renegotiated through strategies aimed at preserving agency and social identity.

### Central process: Maintaining and rebuilding the social self

The analysis revealed a central process through which living with chronic spinal pain becomes organised around a continuous negotiation of the social self. This process, which we name *maintaining and rebuilding the social self*, describes how the social environment shapes, over time, the cognitive and perceptual processes through which chronic pain is lived, interpreted and acted upon. Figure 2 provides an overview of the dynamic relationships between the components of this process.

**Figure 2.**
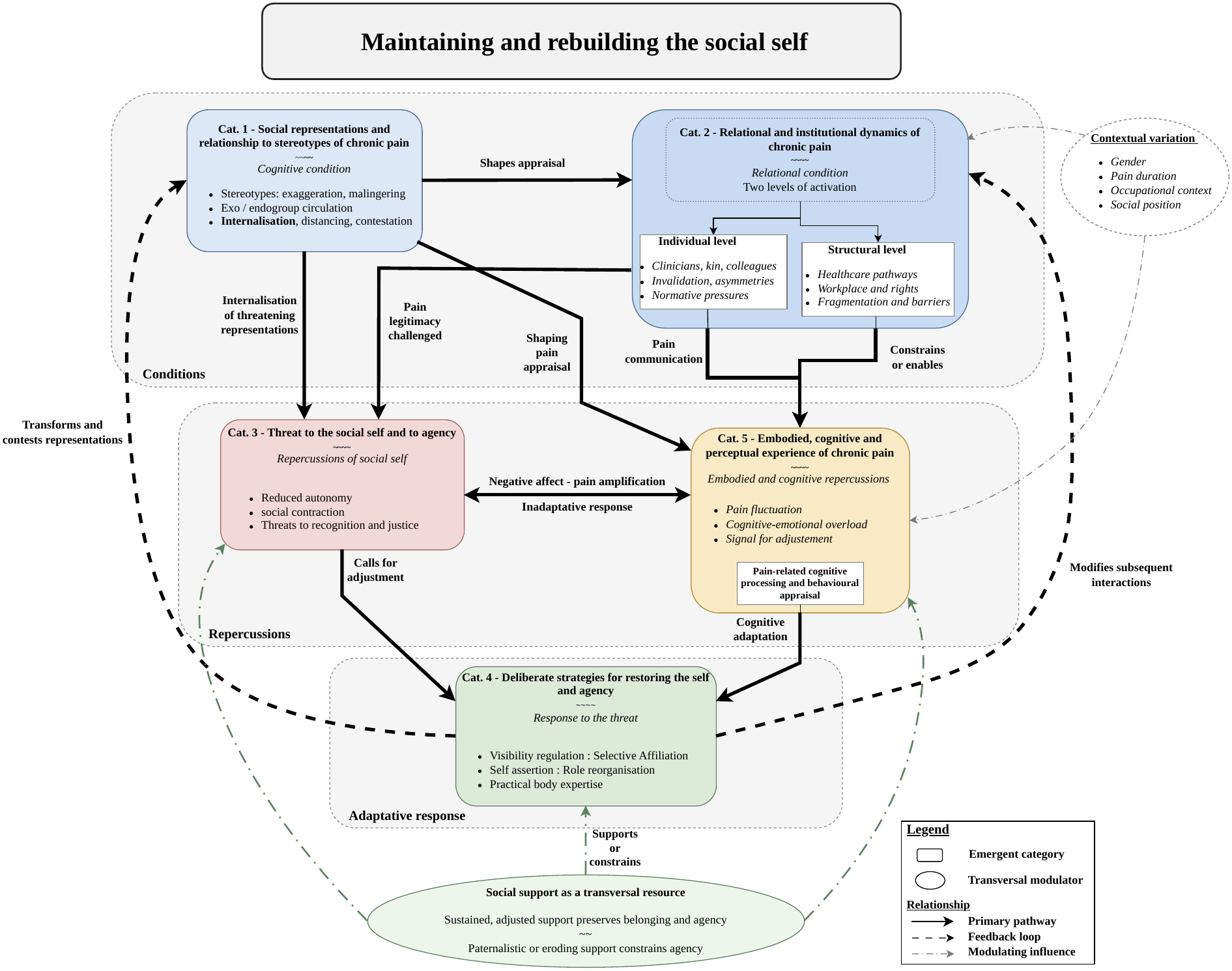
Theoretical model of the central process: Maintaining and rebuilding the social self. The figure presents the theoretical model emerging from the qualitative analysis and informed by constructivist Grounded Theory principles. The model illustrates the dynamic relationships between the five categories and the central process of maintaining and rebuilding the social self. Labels along the arrows identify the key mechanisms linking categories. Solid arrows represent the primary pathway, dashed arrows indicate feedback loops, and dotted arrows represent contextual or social modulation of the process.

The central process unfolds across three analytically distinct components. Social representations and interpersonal interactions (Categories 1 and 2) create the conditions under which the social self becomes threatened. These repercussions (Categories 3 and 5), in turn, call for adaptive responses through which participants attempt to restore agency (Category 4). As participants’ experiences evolve, changes occurring in one component also reshape earlier representations and future interactions. Social support operates across these components rather than as a distinct stage of the process.

Social representations of chronic pain, including stereotypes of exaggeration, moral weakness and illegitimacy, provided interpretive frameworks through which participants approached social interactions. These representations shaped what participants anticipated from others, what they concealed, and how they interpreted others’ responses. These expectations shaped subsequent interactions and were in turn reinforced or revised by experience. The corpus showed a divergence, with lived experience prompting revision for some participants, while others reproduced these representations despite their own pain. What this divergence reveals is that the social self was already being negotiated at the level of representation, before any direct interaction took place. Prior expectations about how others would respond were also reflected in how participants attended to and interpreted their own bodily experience, shaping the cognitive context in which pain signals were processed and given meaning (see figure 2, arrow from Category 1 to Category 5: *shaping pain appraisal*).

These representational frameworks became particularly visible through concrete interactions. Whether in clinical encounters, workplaces or close relationships, what was at stake was the recognition of the person’s pain and, by extension, of the person themselves as a credible actor in their own experience. Whether deliberate or more automatic, delegitimation extended beyond questioning pain itself to challenging the standing of the person reporting it. The passively undergone consequences were an alteration of self-image: a biographical rupture, a sense that one’s own perceptual experience had become subject to others’ authority, a diminished sense of social legitimacy and control. These experiences also shaped how participants approached subsequent interactions, often expecting misunderstanding, disbelief or the need to justify themselves before being recognised as legitimate. Social support ran across all stages of this process rather than constituting a distinct phase (see figure 2, *social support as a transversal resource*). When sustained and well-calibrated, it could buffer this erosion. When it receded over time or tipped into paternalism, it reproduced, in a more benevolent register, the same threat to autonomy that delegitimation posed more overtly.

Participants did not remain passive, however. They reengaged the negotiation from their own side. The deliberate strategies they deployed, interoceptive monitoring, the management of pain’s social visibility, self-assertion against medical and social assignations, and selective affiliation with peers, were oriented not towards pain relief but towards recovering perceived control over their experience and reclaiming a social identity that chronic pain had placed in question. Peer contact carried a particular consequence: it could revise the very representational frameworks that had framed the process from the outset, closing a feedback loop between adaptive responses and cognitive expectations (see figure 2, arrow from Category 4 to Category 1:*transforms and contests representations*). The form and intensity of these strategies varied with contextual factors such as professional status, gender, pain duration and area of residence, which appeared to shape how these processes were experienced, particularly the social pressures participants faced and the resources available to them.

Running through all of this was an experiential dimension that the preceding account risks obscuring. Pain occupied attentional resources, its perceived intensity shaped by emotional states and social interactions alike. The cognitive and perceptual experience of chronic pain was shaped by social interactions and expectations, and in turn influenced how participants interpreted and responded to subsequent situations. What participants had come to expect shaped what they noticed in their bodies. When interactions threatened their standing, the affective weight of pain signals shifted (see figure 2, arrow from Category 5 to Category 3: *negative affect and pain amplification*). And as strategies took hold, how participants read both their own experience and the people around them gradually changed. The social self that participants worked to maintain and rebuild was at once biographical, relational and embodied.

## Discussion

Thirty people living with chronic spinal pain described an experience that extended beyond pain intensity or bodily limitation, and was shaped by the social contexts in which pain had to be recognised and managed. Using an analysis informed by constructivist grounded theory, the analysis led to a model in which social representations, relational dynamics, and cognitive and perceptual processes were closely connected. Together, these dimensions formed a continuous process through which the social self was maintained and rebuilt. By social self we mean the representation of oneself as constructed and continuously negotiated through social interactions (Karos et al., 2018; Markus and Wurf, 1987), distinct from agency (Bandura, 2001), which refers to the perceived capacity to act upon one’s situation. Social experiences reshaped cognitive interpretations, behavioural adjustments, and subsequent expectations over time.

One of the most unexpected findings concerned what happened within the group of people living with chronic pain. Approximately half the participants reproduced, without apparent revision, representations of chronic pain that one might expect to find in the general population. Judgements about simulation, lack of willpower, or inadequate effort were directed at other people with chronic pain. The literature on stigma in chronic pain has focused primarily on interactions between patients and healthcare professionals (Perugino et al., 2022; De Ruddere et al., 2013; De Ruddere and Craig, 2016), and this focus has left largely unexamined what happens within the group. People living with chronic pain can themselves reproduce representations that threaten recognition of chronic pain. Pryma (2017) has described something analogous in the context of contested illness: the implicit moral community that distinguishes legitimate sufferers from malingerers does not operate only from outside. Those inside the community apply it too, often against each other.

Participants also associated legitimacy with specific characteristics. These characteristics can be considered in relation to the dimensions of warmth and competence described in the Stereotype Content Model. Groups associated with disability are commonly positioned within the pity quadrant, characterised by relatively high warmth but low perceived competence (Fiske, 2018). The intragroup prototype in our data only partially matched this pattern. The person with chronic pain was described as organised and capable, with an emphasis on continuing to function despite pain. Restraint in expressing pain was also part of this image and may reflect broader cultural stereotypes of stoicism, which coexist with stereotypes of exaggeration and weakness (Zajacova et al., 2021). Participants appeared to associate legitimacy with competence and restraint, using these criteria to distinguish between people they regarded as truly in pain and those they perceived as exaggerating. Froud et al. (2014) described a related tension: continuing to meet social and occupational expectations despite pain could demonstrate competence. At the same time, this could make the pain itself less credible to others.

A second contribution concerns the recurrent burden of legitimising pain across interpersonal, organisational and institutional settings. Participants described having to make their pain credible not only in healthcare encounters, but also in relationships with relatives, colleagues and supervisors, and within workplace, welfare and administrative systems. At times, the reality or severity of pain was questioned explicitly. At others, social representations and social categorisation processes shaped interactions without deliberate intent. In both cases, participants were repeatedly required to explain, demonstrate or justify limitations that were not readily visible. Chronic pain had to be managed while also being continually rendered socially credible. This recurrent demand for proof had consequences across multiple dimensions of participants’ experience. It contributed to cognitive and emotional overload, complicated decisions about when and how to communicate pain, and intensified threats to the social self. This pattern is consistent with the account of pain as a threat to social needs proposed by Karos et al. (2018), particularly recognition, belonging and autonomy. While some episodes involved deliberate questioning of pain’s reality, others reflected automatic stereotype activation operating without conscious intent. This distinction has direct implications for how rehabilitation training addresses clinician bias (Amodio, 2014).

Social support appeared as a context-dependent and continually negotiated influence rather than a stable resource. Several participants described how those around them gradually stopped accounting for the persistence of their pain, not because it had resolved, but because its continuity had become, for others, unremarkable. Depending on its form and timing, it could buffer threats to the social self or facilitate the restoration of agency. When support took paternalistic forms, it could also reduce participants’ perceived autonomy, although through a different relational process from explicit delegitimation.

One implication of the proposed model is that social representations appear to influence behaviour through the cognitive and perceptual processing of social situations. These interpretations shaped how participants evaluated their own possibilities for action and oriented their efforts to restore the social self. Over time, these efforts and their consequences reshaped future expectations and social representations. Peer contact, in particular, could revise the very representations that had structured the process from the outset, closing a feedback loop and supporting a recursive rather than sequential model. For rehabilitation practice, this suggests that interventions targeting peer learning and social affiliation may not simply provide emotional support, but may act on the representational frameworks through which people with chronic pain interpret both their own experience and that of others. More broadly, the social interactions documented here appear to constitute an important context in which the sources of self-efficacy (Bandura, 1997) are produced, sustained or depleted, an observation that may guide both assessment and intervention in chronic spinal pain rehabilitation.

The present findings also extend the motivational account of pain as a threat to the social self proposed by Karos et al. (2018), by showing empirically how these threats arise and accumulate across everyday social contexts. Rather than representing a simple consequence of chronic pain, maintaining and rebuilding the social self emerges as an ongoing cognitive and relational process through which people continually negotiate recognition, legitimacy and agency. The proposed model also has implications for how chronic pain is recognised and legitimised within clinical practice. Vollert et al. (2026) argue that no biological measure can prove or disprove another person’s pain, and that self-report remains the only indispensable element of clinical assessment. Woo (2026) extends this position by noting that, for many people living with chronic pain, the central problem is not excessive reliance on measurement but persistent failures of recognition, as stereotypes, stigma, and unequal distributions of clinical authority shape whose suffering is acknowledged and whose is not. Coninx and Stilwell (2021) reach a compatible conclusion from a different starting point, arguing that because pain is irreducibly subjective, the social conditions under which it is communicated are not peripheral to clinical assessment but central to it. The present findings provide an experiential account of what this means in practice. Participants described how the absence of a visible biological explanation created a recurring space for doubt, not only in clinical encounters but across workplaces, administrative systems, and close relationships. In each context, uncertainty about pain could become uncertainty about the person reporting it, calling into question the legitimacy of their experience and, consequently, the need for recognition and accommodation. Several participants described anticipating disbelief before seeking care, and at times choosing not to express the full extent of their pain as a result. These accounts show how an epistemological problem, the irreducibility of subjective pain experience to any external criterion, becomes an everyday burden through which the social self is continually negotiated.

### Limitations

Initial and focused coding was carried out primarily by a single researcher, with a second coder involved only for an initial subsample. Although this is consistent with constructivist grounded theory principles, which do not require coding consensus as a marker of objectivity (Charmaz and Thornberg, 2021), it limits the extent to which analytic decisions could be independently traced across the full corpus.

Theoretical saturation was not formally demonstrated through iterative sampling. Data collection and analysis were conducted sequentially, and the sample size was defined in advance (Malterud et al., 2016), but this should not be interpreted as evidence of theoretical saturation in the strict grounded theory sense.

Participants were recruited through tertiary pain assessment and treatment centres. Their accounts may therefore reflect the experiences of people with long, complex healthcare trajectories and repeated encounters with clinical and institutional systems. The model may be less directly transferable to people with chronic spinal pain who are managed in primary care, who do not seek care, or whose pain has not led to prolonged institutional contact.

The study was conducted in France, within a specific healthcare and social protection context. Similar social and relational processes may arise in other settings where the legitimacy of chronic pain is contested, but the forms they take are likely to depend on local institutional and cultural conditions.

## Conclusion

Chronic spinal pain is experienced within social contexts in which it may need to be explained, justified, and defended. The model proposed here describes how this process shapes cognitive expectations, threatens the social self, and leads people to develop strategies for preserving agency and social identity. The contribution of this study is to show how social representations, interactions, and responses become linked over time in the ongoing negotiation of pain legitimacy.

Recognising these findings may help rehabilitation professionals to understand how people continually negotiate the credibility of their pain and, through this, maintain and rebuild the social self. Future longitudinal and mixed-methods studies should examine how these processes evolve over time and whether interventions targeting social interactions, peer learning, and pain legitimacy can improve rehabilitation outcomes.

## Author Contributions

The authors have adopted the CRediT (Contributor Roles Taxonomy) system to specify individual contributions:

- **YS:** Conceptualization, Methodology, Project administration, Funding acquisition, Investigation, Formal analysis, Data curation, Writing – original draft, Writing – review & editing.
- **AC:** Investigation, Data curation.
- **DM:** Data curation, Formal analysis.
- **JBVH:** Methodology, Formal analysis, Writing – review & editing, Supervision.
- **EC:** Conceptualization, Methodology, Project administration, Resources, Formal analysis, Writing – review & editing, Supervision.

All authors have read and agreed to the published version of the manuscript.

## Acknowledgements

The authors would like to express their sincere gratitude to:

- All the **participants** and **healthcare professionals** who participated in this study and generously shared their time and experiences.
- The staff of the **CETDs** (Centres d’Évaluation et de Traitement de la Douleur) for their warm welcome and for facilitating participant recruitment.
- **Nicolas Naiditch, PhD** (Sociology Researcher) and **Evelyne Lasserre, PhD** (Lecturer in Anthropology, University of Lyon 1) for their valuable expertise and guidance in the methodological design of the study and the development of the interview guide.

## Additional information

### Data availability statement

The data that support the findings of this study are not publicly available due to privacy and ethical restrictions concerning participant information. Study materials, including the interview guide, codebook, original French quotations, and high-resolution analytic diagrams, are available on Zenodo: https://doi.org/10.5281/zenodo.23963293.

### Declaration of generative AI use

The authors used an AI language model (Anthropic Claude Sonnet 4.6) to assist with English translation and sentence reformulation during the preparation of this manuscript. All intellectual content was written and reviewed by the authors, who take full responsibility for the final version of the publication.

